# Exploring the Healthy Behaviors of Nigerians during the COVID-19 Pandemic

**DOI:** 10.1101/2020.11.25.20235457

**Authors:** Ifeanyichukwu Meek Eyisi

**Affiliations:** Department of Psychology, Covenant University, Ota, Ogun State, Nigeria

**Keywords:** COVID-19, Healthy behavior, Nigerians, Thematic analysis, Qualitative study

## Abstract

Healthy behaviors remain important for staying safe during the coronavirus disease 2019 (COVID-19) pandemic. This study, therefore, explored the healthy behaviors of Nigerians during the COVID-19 pandemic and the impact of COVID-19 related news on healthy behaviors. Thirty-three (17 females and 16 males) participants from the general Nigerian population with age range of 23-64 years were recruited via social media using the snowball technique. Responses were elicited using semi-structured questions and subjected to thematic analysis. The healthy behaviors identified included; “social distancing”, “changes in nutrition”, “hand washing or sanitizing”, “exercise”, “increased vigilance from those with comorbidities”, and “use of facemask”. In another analysis, the impacts of COVID-19 related news on healthy behaviors were; “behavior modification”, “anxious impacts”, and “fake news about COVID-19 caused people to stop listening to COVID-19 related news”. Findings generated practical implications for enhancing healthy behaviors during the COVID-19 pandemic. The role of the media in strengthening healthy behaviors during the pandemic was also highlighted.

## INTRODUCTION

Healthy behaviors remain a very crucial aspect of every illness. Healthy behaviors describe the actions we embark on to prevent diseases or illnesses (Haighton, et al. 2013). What this means is that the kind of behavior we practice can either be a protective or risk factor for us to contract illnesses. Literature suggests that more than 50% of mortality related to illness is a function of the behavior (McKeown, 1979). This is suggestive that behaviors may be doing more harm than an illness itself. More current literature supports this assertion, for instance, Klein, et al. (2015) have linked cancer prevention and control with behaviors. The same argument was reinforced by Kanera, et al. (2016) who established a significant association between healthy behaviors and cancer illness among early cancer survivors. In another clinical population of people living with HIV/AIDS (PLWHA), Radfar, et al. (2014) determined that healthy behaviors such as protective sexual intercourse was significantly interactive with HIV transmission among PLWHAs in Iran. In Nigeria, Ogueji and Okoloba (2020) reported that unhealthy behaviors such as suicidal ideation was associated with poor coping among newly diagnosed people with HIV/AIDS in Ibadan, Nigeria.

Despite the above implications that healthy behaviors have on disease contraction, there is currently limited information that is available about the healthy behaviors that people are practicing during the novel COVID-19 pandemic. Related studies often focused on the mental or physical health impacts of the COVID-19 pandemic on people from various countries. For instance, Okoloba et al. (2020) determined the presence of anxiety and depression in a multinational sample (e.g., participants from Nigeria, the United Kingdom, the Netherlands, the United States of America, United Arab Emirates, Singapore, etc.,) who were affected by the experience of the COVID-19 pandemic. In the same study, it was recurrently found that most participants experienced drastic changes in their lifestyles due to the COVID-19 pandemic, and most found it uneasy coping with the changes. In a similar study, Olaseni, et al. (2020) reported the presence of anxiety and psychological distress among Nigerians amid the COVID-19 pandemic. Some other studies have also found the presence of depression, post-traumatic stress disorder (PTSD), and sleep problems among people during the COVID-19 pandemic (Huang & Zhao, 2020; Gao, et al. 2020; Ozdin & Ozdin, 2020).

Serafini et al. (2020) submitted from a comprehensive literature review that among the impacts of the COVID-19 pandemic were stress, anxiety, depression, frustration, and uncertainty. In addition, the same findings showed adverse impacts of quarantine and adverse impacts from the protective measures implemented to control the spread of the COVID-19 pandemic. Such adverse impacts included increased anxiety and obsessive and compulsive disorder (OCD) from the general public. An implication of these findings suggests the need to ensure that researches supporting the adoption of healthy behaviors in the general public are encouraged and thus strengthened.

In another study, Zhang and Ma (2020) established that the impacts of COVID-19 pandemic included mild stress, helplessness, horrific and apprehensive concerns. Furthermore, suicidal thoughts were also found among study participants. Although the study provided some insights into the mental health impacts of the COVID-19 pandemic, the authors suggested that further studies or a different research method will be helpful to establish if the same findings will resurface. In a different study, Cao et al. (2020) found that among vulnerable students, the major impacts of the COVID-19 pandemic included anxiety symptoms, concerns about academic delay, economic effects and the impacts on daily living such as restricted movements.

Kim et al. (2020) in South Africa found that about 14.5% of adults were at risk of depression during the pandemic. In that same study, it was qualitatively found that experiences of anxiety, financial insecurity, fear of infection, and rumination were also found among participants. Fegert et al. (2020) submitted that the mental health threats associated with the COVID-19 pandemic is adversely affecting the mental health of children and adolescents. The researchers also predicted that the COVID-19 associated mental health risks will disproportionately hit marginalized and disadvantaged children compared to children from normal/healthy homes. Therefore, research is needed to assess the implications of policies and interventions for children.

Prior to the COVID-19 pandemic incidence, there were infectious disease outbreaks such as the severe acute respiratory syndrome coronavirus (SARS-COV). In terms of the health impacts from that outbreak, literature has found issues of stigmatization, anxiety, and stress as common reports presented by patients with the SARS-COV. In another research. Nickell et al. (2004) found that emotional distress was commonly reported among people working with patients of SARS-COV. Almost similarly, Sprang and Silman, (2013) embarked on a comparative study and found that anxiety, depression, PTSD, and alcohol abuse was common among persons quarantined during the SARS-COV.

Wu, et al. (2008) study reinforced this by establishing that alcohol misuse was common after three years of the SARS outbreak, with alcohol dependency been positively correlated with having been quarantined and self-isolated. In a different study, Almutairi et al, (2015) found that the SARS-COV brought about effects such as high levels of concern, and agitation from the general public. Overall, the above literature is suggestive that the COVID-19 pandemic as well as similar infectious disease outbreaks often limits normal functioning in people in the society, and this poses threats to the physical and mental health of people in the society.

Given the mental and physical health impacts that are associated with the COVID-19 pandemic, it is thus worthwhile to explore the healthy behaviors that people are practicing during the pandemic. This is imperative to inform appropriate interventions that strengthen peoples’ healthy behaviors and reduce their chances of contracting the COVID-19 infection. Therefore, the following research questions were thus generated based on the review of the above literature:

1. What healthy behaviors are people practicing during the COVID-19 pandemic?
2. How has the COVID-19 related news from the media affected the healthy behaviors of people during the COVID-19 pandemic?

## METHOD AND DATA

### Design

This was a qualitative semi-structured research design that explored the healthy behaviors that people are practicing during the COVID-19 pandemic. The design was adopted because it permitted the participants to freely express themselves. Thus, giving voices to the participants, in order for the researcher to truly understand the experiences and needs of the participants.

### Sampling and Participants

A snowball sampling technique was deployed between July and August 2020. Participants were eligible if they were 18 years or above, resided in Nigeria, and had internet access. The researcher sent the survey link via WhatsApp, and Facebook. The researcher also encouraged participants to send out the survey link to their counterparts who were willing to participate in the study. Seventeen females and 16 males with age range of 23-64 years participated in this study. All participants were Nigerians. One has senior secondary school certificate examination (SSCE) as the highest education attained, while 32 had acquired education from tertiary institutions.

### Materials

This study designed two (2) qualitative questions into a single survey link using the Google forms. The survey also elicited demographic information from participants, which included; gender, age, country of residence, and educational qualification. The 2 questions asked were: (1) “Describe the healthy behaviors that you are adopting during the COVID-19 pandemic” (2) “Do you watch news about COVID-19? If yes, how has the news about COVID-19 affected your healthy behavior”.

### Procedure

The research was approved by the Research Ethics committee of Covenant University, Ota, Ogun State, Nigeria. The researcher sent out the survey link to WhatsApp groups and Facebook in order to recruit participants. The purpose of the study was highlighted in the survey link, and an online consent form was inclusive. All participants maintained anonymous identities so that they could be sincere about their responses. Participation in the study was voluntary and confidentiality of information was assured to all participants. Participants who filled the survey were encouraged forward the survey to their counterparts.

While the data collection was ongoing, the researcher monitored the number participants thus gathered, and when the researcher was satisfied with the detailed response of participants, he ended the data collection process, and proceeded to data analysis. Data was analyzed using thematic analysis for the purpose of extracting relevant themes from the responses given by respondents. The author read the responses of respondents in order to derive common themes, and responses were placed under each theme that was most appropriate for them. Participants were given feedback from the data analysis, and themes were reworked where necessary.

## RESULTS

Qualitative question 1: Describe the healthy behaviors that you are adopting during the COVID-19 pandemic.

### Social distancing

This was the first healthy behavior identified, as 20 participants mentioned that they have been observing social distancing. Quoted example is below:

> *“Since the COVID-19 pandemic started, I have been observing 2 meters distancing…” (Male, 50’s)*.

A female also said that:

> *“I go out only when necessary as few times per week. Usually once, sometimes twice, and only visiting those in my ‘bubble’…” (Female, 40’s)*.

### Changes in nutrition

This was the second theme found. Participants said that they have resorted to eating healthier.

> *“I have been eating healthier foods thus far to stay safe…” (Female, 50’s)*

A female reinforced this by expressing how she had cut down on snacks.

> *“Cutting down on snacking, and trying to drink plenty of water…” (Female, 50’s)*.

Male participants also reinforced this theme by expressing their increased consumption of vitamins and vegetables.

> *“Taking vitamins to boost my immune system” (Male, 20’s)*.
>
> *“We have increased our intake of vegetables as a healthy measure…” (Male, 30’s)*

### Hand washing or sanitizing

They mentioned that they were regularly washing or sanitizing their hands as part of their healthy behaviors.

> *“…Washing my hands thoroughly with soap and water and increased use of sanitizers…” (Female, 20’s)*.
>
> *“I regularly use my hand sanitizer after counting money or touching places outside my home…” (Female, 40’s)*.

### Exercise

A hand-full mentioned that frequent exercises have been a healthy measure for them.

> *“I am having my daily walk in the morning as part of keeping healthy…” (Female, 50’s)*.
>
> *“I always do exercise to stay healthy during the pandemic…” (Male, 40’s)*

### Increased vigilance from those with comorbidities

Those with a comorbidity said that they have become extra vigilant due to the pandemic.

> *“My major healthy behavior is that I am extra vigilant as I am high-risk with comorbidities” (Male, 50’s)*.

### Use of facemask

A hand-full reported the use of facemask as their healthy behaviors. For example;

> *“…Putting on face mask when outside is a major healthy behavior that I follow” (Female, 20’s)*.

Qualitative question 2: Do you watch news about COVID-19? If yes, how has the news about COVID-19 affected your healthy behavior?

### Behavior modification

This was the first theme. Over 25 participants mentioned that information from COVID-19 related news has brought about behavioral modifications for them. Examples are thus below:

> *“The news has modified my behavior, and I have changed to conform to the COVID-19 rules and regulations when in public*.*” (Male, 50’s)*.

They also mentioned that watching news increased their attention on the seriousness of the pandemic.

> *“Listening to the news made me to take COVID-19 seriously, and I started social distancing and work from home before the official government notification to stay home*.*” (Female, 30’s)*.
>
> *“Yes, I do watch news about COVID-19 and my health behavior changed by improving my personal hygiene*.*” (Female, 40’s)*.

### Anxious impacts

They self-reported that watching news about COVID-19 had anxious impacts on them.

> *“Watching the news is worrying as I’m a mother and I worry about my own health as well as that of my family” (Female, 50’s)*.
>
> *“Yes, I had anxiety after watching the news but after a while I adjusted” (Female, 30’s)*.

### Fake news about COVID-19 caused people to stop listening to COVID-19 related news

They also mentioned that fake news about COVID-19 made them to stop watching COVID-19 related news.

> *“I no longer watch the news about COVID-19! Information reported is inaccurate! (Female, 50’s)*.

## DISCUSSION AND IMPLICATION

This qualitative study explored the healthy behaviors of Nigerians during the COVID-19 pandemic. Two questions were asked. The first question asked participants to describe the healthy behaviors that they were adopting during COVID-19 pandemic. Conducting thematic analysis, the following themes emerged from the first question. They included; social distancing, changes in nutrition, hand washing or sanitizing, exercise, increased vigilance from those with comorbidities, and use of facemask.

Second question asked participants if they watch COVID-19 related news, and the impact that information from the news had on their healthy behaviors. Conducting thematic analysis, the following themes emerged from the second question. They included; behavior modification, anxious impacts, and fake news about COVID-19 caused people to stop listening to COVID-19 related news.

One implication of the above findings mirrors that the COVID-19 pandemic has brought about changes in daily living for Nigerians that are affected by it. Some of the changes can be said to be triggered by the fear of contracting the disease, or the preventive/control measures that are implemented to protect the public health. An additional implication suggests that the COVID-19 related news from the media can have the potential to strengthen the practice of healthy behaviors if the news contents align with the needs of the public.

The findings from this qualitative study aligned with the literature where identical themes have emerged from the impacts of the COVID-19 pandemic (Olaseni, et al. 2020; Okoloba, et al. 2020; Ozdin & Ozdin, 2020). On the bases of findings in the current study, the researcher recommends that the themes that emerged in this study should inform the development of interventions that are aimed at strengthening the practice of healthy behaviors during the COVID-19 pandemic. The media should also be used as an agent to strengthen the practice healthy behaviors, rather than an agent for spreading COVID-19-induced fear.

This study has some limitations that must be acknowledged. First, the method of data collection did not accommodate digitally excluded people whose healthy behaviors during the COVID-19 pandemic may be particularly important to understand. Second, the research design and sampling technique can compromise the generalization of findings. Another limitation of the study was that the participants self-reported anxious impacts from COVID-19 related news, and this research did not include measures of anxiety in data collection. It is, consequently, not possible to explain how anxious participants were. Lastly, the participants were recruited from Nigeria, and it is not certain that the findings from this research represent the healthy behaviors of people in the western worlds that were particularly hit by the global pandemic. Nonetheless, the findings from this research contributes to the growing body of knowledge on the COVID-19 pandemic.

This study concluded that Nigerian participants in this study were practicing healthy behaviors to stay safe during the COVID-19 pandemic, and COVID-19 related news from the media was positively and negatively affecting the practice of healthy behaviors among them. This research can be furthered by exploring the barriers to practicing healthy behaviors during the COVID-19 pandemic.

## Data Availability

Data is available from the author on reasonable request.

## ACKNOWLEDGEMENT

Special thanks to the participants.

## ETHICAL CONSIDERATIONS

The research was approved by the Research Ethics committee of Covenant University, Ota, Ogun State, Nigeria. This study conformed to the Helsinki 1964 ethical declaration, its later amendment, or a comparable standard. Online consent form was used to elicit consent from participants.

## DATA AVAILABILITY STATEMENT

Data is available from the author on reasonable request.

